# ADHD symptom trajectories and brain morphometry: A longitudinal analysis

**DOI:** 10.64898/2026.04.07.26350043

**Authors:** Aylin Mehren, Jill Kessen, Aleksandra M. Sobolewska, Daan van Rooij, Jaap Oosterlaan, Catharina A. Hartman, Pieter J. Hoekstra, Marjolein Luman, Anderson M. Winkler, Barbara Franke, Jan K. Buitelaar, Emma Sprooten

## Abstract

**Objective:** While ADHD symptoms often decline from childhood into adulthood, the underlying neurobiological mechanisms, such as altered brain maturation or neural reorganization, remain incompletely understood. This study investigated how grey matter development relates to ADHD symptom trajectories into adulthood.

**Method:** We analyzed data of individuals with ADHD and controls from the longitudinal Dutch NeuroIMAGE cohort, utilizing dimensional ADHD symptom scores (Conners Parent Rating Scale) from three waves and T1-weighted structural MRI scans from the final two waves. Using General Linear Models with permutation-based inference, we examined: 1) cross-sectional associations between ADHD symptoms and vertex-wise cortical thickness and surface area, and subcortical volumes at Wave 1 (n=765, mean age =16.95 years); and 2) longitudinal associations between symptom progression and brain morphometric changes (Wave 0 to 1: n=644, mean age=11.55–17.24 years; Wave 1 to 2: n=149, mean age=16.45–20.11 years).

**Results:** Cross-sectionally, at Wave 1, more ADHD symptoms were related to widespread reductions in surface area, most prominently in the frontal cortex, and smaller volumes of the cerebellum, amygdala, and hippocampus. Longitudinally, symptom improvement from Wave 1 to Wave 2 was associated with stronger reductions in surface area, particularly in prefrontal and occipital regions, and with more pronounced cortical thinning across multiple brain regions.

**Conclusion:** These findings suggest an association between symptom trajectories and structural brain changes, indicating that clinical improvement in ADHD behaviors might coincide with ongoing neural refinement during the transition to adulthood.

## Introduction

Attention-deficit/hyperactivity disorder (ADHD) is a common neurodevelopmental condition with a prevalence of 5.9% in childhood, and is characterized by impairing symptoms of inattention, and/or hyperactivity and impulsivity^1^. The diagnosis persists into adulthood in approximately half of the individuals and an additional proportion continues to experience subclinical symptoms^1^. In general, ADHD symptoms tend to decrease from childhood to adulthood. However, recent evidence highlights a dynamic longitudinal course characterized by diverse patterns of symptom fluctuation, with individuals experiencing shifting periods of remission, subclinical symptoms, persistent impairment, or even worsening of clinical outcomes^2,3^.

ADHD is understood to arise from a complex interplay of genetic, environmental, and neurobiological factors^1^. Neuroimaging studies have demonstrated subtle but wide-spread variations in brain structure and function related to ADHD^4–7^. Notably, meta-analyses and large-scale studies indicate that some neuroanatomical variations are stronger in children compared to adults with ADHD^4,5,8^. Moreover, the neural mechanisms underpinning the diverse developmental and clinical trajectories of ADHD remain incompletely understood and may differ from those associated with the initial manifestation of ADHD.

Neurodevelopmental theories propose several mechanisms to explain ADHD remission versus persistence into adulthood. One model suggests a process of convergence toward neurotypical patterns, often characterized by a delayed but eventual catch-up in cortical maturation. On the other hand, remission may be driven by compensation and neural reorganization^9,10^. Shaw et al.^11^ pioneered support for the idea of "delayed cortical maturation" in children with ADHD. Their longitudinal analysis of cortical growth demonstrated that while cortical thickness developed in a similar manner in children with and without ADHD, those with ADHD attained their maximum cortical thickness at a later age. This delay was observed across many regions of the cortical surface, and in particular in the prefrontal cortex. Subsequent work showed reductions in cortical surface area in frontal, parietal, temporal, and occipital regions related to ADHD in childhood, which converged towards neurotypical averages in young adulthood, but again, individuals with ADHD reached their peak surface area at a later age^12^.

Conversely, some studies suggest that certain brain characteristics associated with ADHD may persist into adulthood. Using independent component analysis within a multivariate approach, Cupertino et al.^13^ identified an ADHD-related component reflecting reduced volume in the frontal lobes, striatum, and interconnecting white matter that was independent of age, appearing consistently in both childhood-to-adolescent and adult replication samples, albeit in cross-sectional data. Those findings together suggest that some brain differences might disappear with age, while others might stay stable. Similarly, investigations into white matter development indicate that distinct neurodevelopmental mechanisms, such as normalization and compensation, can occur in parallel rather than being mutually exclusive, highlighting the complexity of brain changes in ADHD^14,15^.

Despite these valuable insights, comprehensive longitudinal studies investigating the neurobiological mechanisms associated with the developmental course of ADHD remain limited, often constrained by small sample sizes or restricted age ranges. Addressing this critical gap, the current study aimed to investigate how grey matter development relates to ADHD symptom trajectories from childhood into adulthood. Moving beyond region-of-interest and atlas-based analyses, we employed data-driven whole-brain vertex-wise analyses of cortical thickness and surface area, as well as subcortical volumes. Our analyses used data from the Dutch NeuroIMAGE study^16–18^, a large-scale longitudinal project following individuals with and without ADHD from childhood to young adulthood. The study consists of three waves of clinical data, with brain imaging collected during the final two waves.

Adopting a dimensional perspective, we employed a stepwise approach to examine ADHD as a continuous trait. First, we examined if grey matter indices are related to ADHD symptom scores at Wave 1 (cross-sectional analysis). Here, we hypothesized subtle reductions in vertex-wise cortical thickness, surface area, and subcortical volumes in several regions (especially prefrontal cortex and striatum/basal ganglia) in relation to higher continuous symptom scores. Second, we longitudinally investigated whether different ADHD symptom trajectories across development are associated with distinct patterns of grey matter development (from Wave 0 to Wave 1 and from Wave 1 to Wave 2). We hypothesized that symptom reductions are associated with two complementary patterns: (a) normalization/convergence, involving an attenuation or catching-up of some initial grey matter reductions; (b) compensation, involving grey matter increases in other brain regions supporting symptom reduction.

## Method

This study was pre-registered on the Open Science Framework (OSF; https://osf.io/4ysjh) prior to data inspection and analysis. The pre-registration includes hypotheses, study design, and planned analyses.

### Participants

Data for this study were drawn from three waves of the Dutch IMAGE (Wave 0) and NeuroIMAGE (Waves 1 and 2) studies, and included clinical and neuroimaging data from individuals with ADHD, their first-degree relatives, and families without ADHD. Details on study designs, inclusion criteria and data collection have been described previously^16–18^. In Waves 0 and 1, data was collected in Nijmegen and Amsterdam and in Wave 2 in Nijmegen. MRI data was only collected in Waves 1 and 2. The studies were approved by the local medical ethics committees, and written informed consent was obtained from all participants and their legal guardians.

Our cross-sectional analyses included all participants from Wave 1 with at least one adequate-quality T1-weighted structural MRI scan and concurrently collected ADHD symptom scores (n=767 before and n=765 after final quality control). The longitudinal analyses included participants with ADHD symptom scores available from at least two waves. In addition, for the analysis from Wave 0 to Wave 1, an adequate-quality T1-weighted scan at Wave 1 was required (n=646). In this analysis, age/birthdate was missing for two participants, so the final sample consisted of n=644 participants. For the analysis from Wave 1 to Wave 2, adequate-quality scans for both waves were prerequisite (n=150 before and n=149 after final quality control). Table 1 shows demographic and clinical characteristics of the final samples.

**Table 1.**
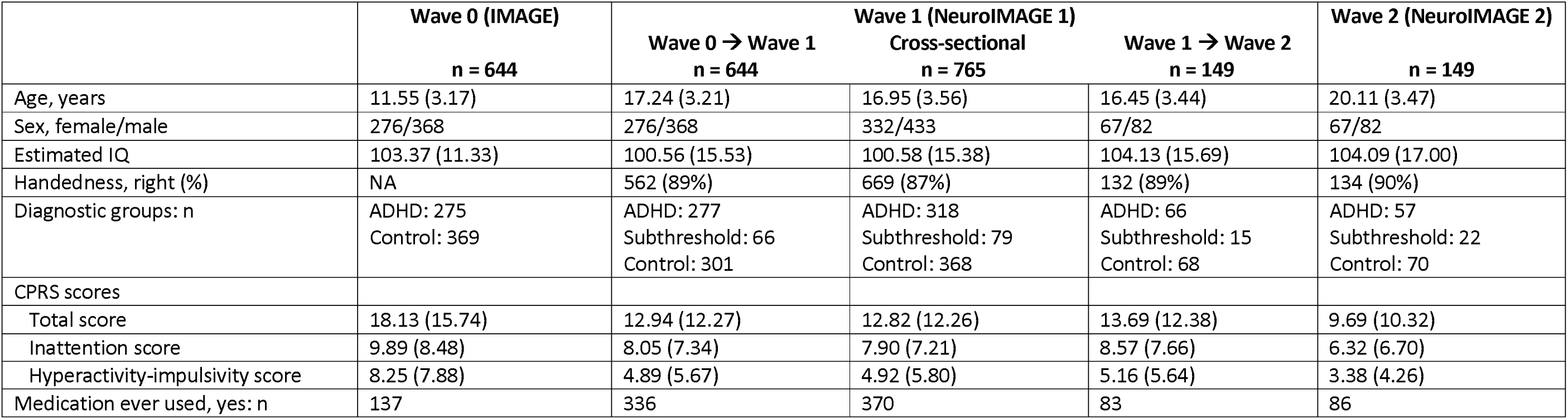
Demographic and clinical characteristics of the sample at each wave. Reported values pertain to all participants in the final sample after final quality control. IQ: estimated using vocabulary and block design subtests of the Wechsler Intelligence Scale for Children or Wechsler Adult Intelligence Scale. CPRS: Conners Parent Rating Scale, total score is the sum of the scores for hyperactivity-impulsivity and inattention. Medication ever used: Whether or not participants had ever taken methylphenidate, atomoxetine or any other ADHD medication.

Our dataset provides a comprehensive view of ADHD symptom severities and their longitudinal trajectories, spanning individuals with a clinical ADHD diagnosis, those with subthreshold symptoms, and unaffected controls. Crucially, diagnoses for some participants demonstrated variability across timepoints. To best capture the dynamic and dimensional nature of ADHD, our main analyses conceptualized ADHD as a continuous trait. This continuous framework has shown sensitivity to diffusion-weighted brain features in previous analyses using the same dataset^19,20^. For comparability with prior studies, we additionally performed an exploratory analysis using diagnostic categories (ADHD vs. non-ADHD, after removing individuals with subthreshold ADHD).

### Clinical measurements

ADHD symptoms were assessed using the Conners Parent Rating Scale (CPRS)^21^. Parents rated 80 items on a 4-point Likert scale (0=“not true at all” to 3=“very much true”). As primary measure we used the raw DSM-IV total ADHD score, encompassing 9 items assessing DSM-IV criteria for inattention and 9 items for DSM-IV hyperactivity-impulsivity symptoms. In secondary analyses, we examined raw DSM-IV scores for inattention and hyperactivity-impulsivity separately. For longitudinal analyses, symptom change was computed as the difference between ADHD scores between the respective waves (CPRS_Wave0–CPRS_Wave1; CPRS_Wave1–CPRS_Wave2), where more positive values indicate greater symptom improvement. Diagnostic categories were determined based on an algorithm published previously^18^, combining a diagnostic interview (K-SADS) with Conners rating scales.

### Structural MRI processing

Details of MRI acquisition are described in Supplement 1. During initial quality control, raw T1-weighted structural images were visually inspected and rated from 1=no distortions to 4=severe distortions^22^. Images with adequate quality (rating of 1 or 2, no or mild distortions) were included for further processing. In cases of two available adequate-quality scans for one participant at a single wave, the scan with highest quality was utilized for subsequent processing.

Cortical reconstruction was performed using FreeSurfer version 7.3.2 (http://surfer.nmr.mgh.harvard.edu/). FreeSurfer is an automated technique to create a surface-based reconstruction of the cortical sheet that uses both intensity and continuity information^23^, with good test–retest reliability across scanner manufacturers^24,25^. For longitudinal analyses, images were processed using the longitudinal FreeSurfer stream to extract reliable volume and thickness estimates^25^.

Post-processing quality control involved visual inspection of all images initially rated with 2 (mild distortions) and inspection of images with more topological defects based on Euler number calculation (>1 SD away from mean). In addition, outliers in brain measures were checked for processing errors and physiological plausibility. For longitudinal analyses, all within-subject template images and final longitudinal images were inspected, as recommended for pediatric populations.

A 10-mm full width at half maximum surface-based smoothing kernel was applied. Cortical thickness was calculated as the closest distance from the gray/white boundary to the gray/CSF boundary at each vertex on the tessellated surface. Surface area was calculated at the geometric middle between the inner and outer cortical surfaces at each vertex, and interpolated for between-subject analyses using a quantity-conserving method^26^. Volumes were calculated for the cerebellum and the following subcortical structures of the left and right hemispheres: nucleus accumbens, caudate nucleus, hippocampus, putamen, amygdala, pallidum, and thalamus. Average cortical thickness and total surface area were calculated across all vertices, and total intracranial volume was estimated based on the linear registration to a standard brain in FreeSurfer.

### Statistical analyses

Statistical analyses were performed using general linear models (GLMs) in Freesurfer to investigate the association between ADHD symptom scores (predictor) and whole-brain vertex-wise cortical thickness, surface area, or subcortical volumes (dependent variables). For cross-sectional analyses, GLMs were constructed to assess the association between ADHD scores and brain morphometric outcomes both at Wave 1. Sex, age, and scanner site were included as covariates. Longitudinal associations were examined in two stages. First, we investigated the association between the change in ADHD scores from Wave 0 to Wave 1 and brain outcomes at Wave 1. Covariates included sex, age at Wave 0, age difference between waves, ADHD score at Wave 0, and scanner site. Second, we examined the association between the change in ADHD scores from Wave 1 to Wave 2 and brain measures at Wave 1, Wave 2, and the change in brain measures between Wave 1 and Wave 2. Covariates included sex, age at Wave 1, age difference between waves, and ADHD score at Wave 1. Figure 1 shows an overview of the longitudinal analyses.

**Figure 1.**
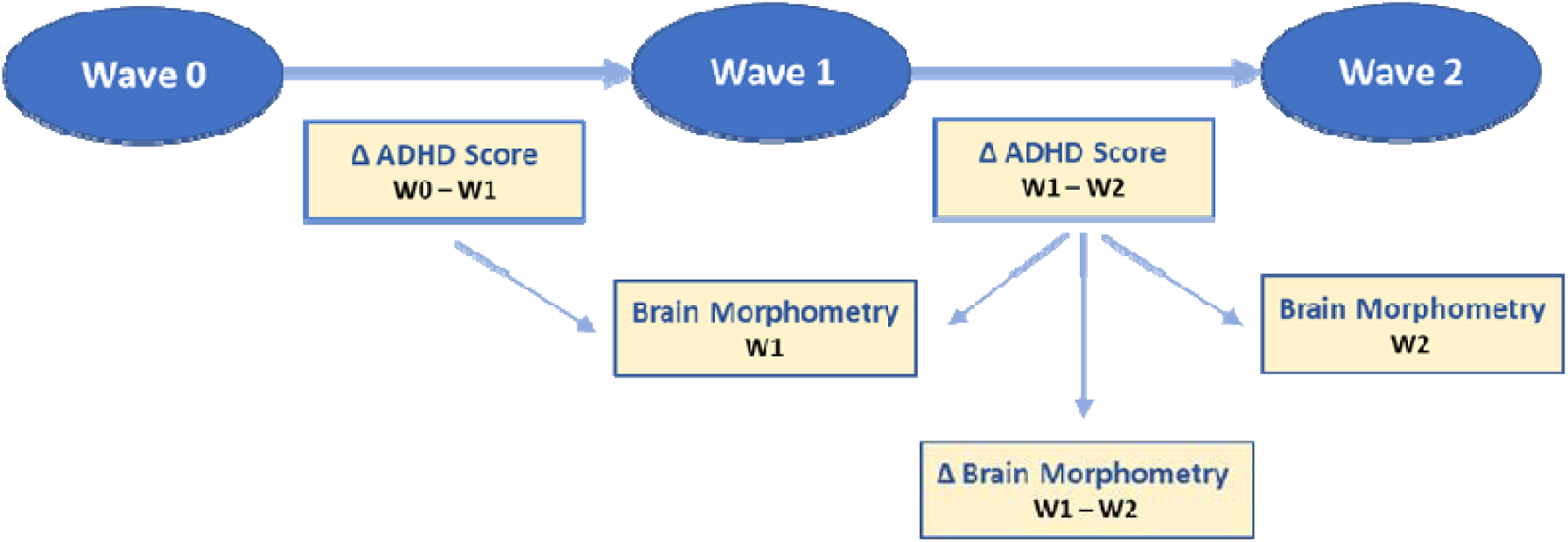
Overview of the longitudinal analyses. For each analysis, brain morphometry outcomes included whole-brain vertex-wise surface area and cortical thickness, and subcortical volumes.

Primary analyses utilized the DSM-IV total ADHD score as predictor, secondary analyses included DSM-IV scores for inattention and hyperactivity-impulsivity separately as predictors. Sensitivity analyses were conducted post-hoc for any significant findings. These involved re-running the GLMs with whole-brain average cortical thickness, total surface area, total intracranial volume, or ADHD medication usage (ever used, yes/no) as additional covariates. Additionally, separate GLMs were conducted with global brain metrics as dependent variables. For significant findings in the cross-sectional analyses, additional GLMs were performed to compare individuals with (n=318) and without (n=368) ADHD at Wave 1, excluding those with subthreshold ADHD (n=79). Group comparisons were not performed for longitudinal analyses due to small sample sizes when considering diagnostic group-combinations across timepoints (see Table 1). To aid interpretation of significant findings in the longitudinal analyses, post-hoc analyses were conducted to examine the association between baseline ADHD scores and brain structural changes, both with and without symptom change as covariate. Additional sensitivity analyses excluding baseline ADHD scores from the primary models were conducted to assess the potential influence of baseline symptom severity on the longitudinal findings.

Permutation-based multiple comparison correction was applied using Permutation Analysis of Linear Models (PALM)^27^ in Matlab. Each model underwent 5000 permutations. To account for the non-independence of data due to sibling relationships and shared familial variance, permutation tests were constrained using multi-level exchangeability blocks, as described by Winkler et al.^28^. This method restricted permutations both at the whole-block level (i.e., between families of the same size) and the within-block level (i.e., within families). Because variances could not be assumed to be homogeneous, variance groups were specified by the exchangeability blocks, and Aspin-Welch’s v-statistics (vstat), which are analogous to t-statistics but account for heteroscedasticity, were calculated. For cortical data, threshold-free cluster enhancement (TFCE) option was used, with Family-wise error rate (FWE)-corrected p<.05 considered statistically significant. For subcortical volumes, false discovery rate (FDR) correction was applied in PALM, with FDR-corrected p<.05 considered statistically significant.

## Results

### Cross-sectional associations between ADHD symptom scores and brain morphometric measures at Wave 1

At Wave 1, total ADHD score was significantly negatively associated with surface area in one large left-hemisphere and one large right-hemisphere cluster, with predominantly small effect sizes within each vertex (left: size=56214.75 mm^2^, p_FWE_<.05; right: size=56961.24 mm^2^, p_FWE_<.05; Figure 2) and overall strongest effects in the frontal cortex. No significant associations were found between total ADHD score and vertex-wise cortical thickness. Furthermore, total ADHD scores were negatively associated with volumes of the left and right cerebellum (left: vstat=-2.97, p_FDR_=.02, df=10.64; right: vstat=-3.52, p_FDR_=.008, df=10.58), hippocampus (left: vstat=-2.64, p_FDR_=.0496, df=10.70; right: vstat=-2.61, p_FDR_=.0496, df=10.54), and amygdala (left: vstat=-3.58, p_FDR_=.008, df=10.65; right: vstat=-3.13, p_FDR_=.02, df=10.78).

**Figure 2.**
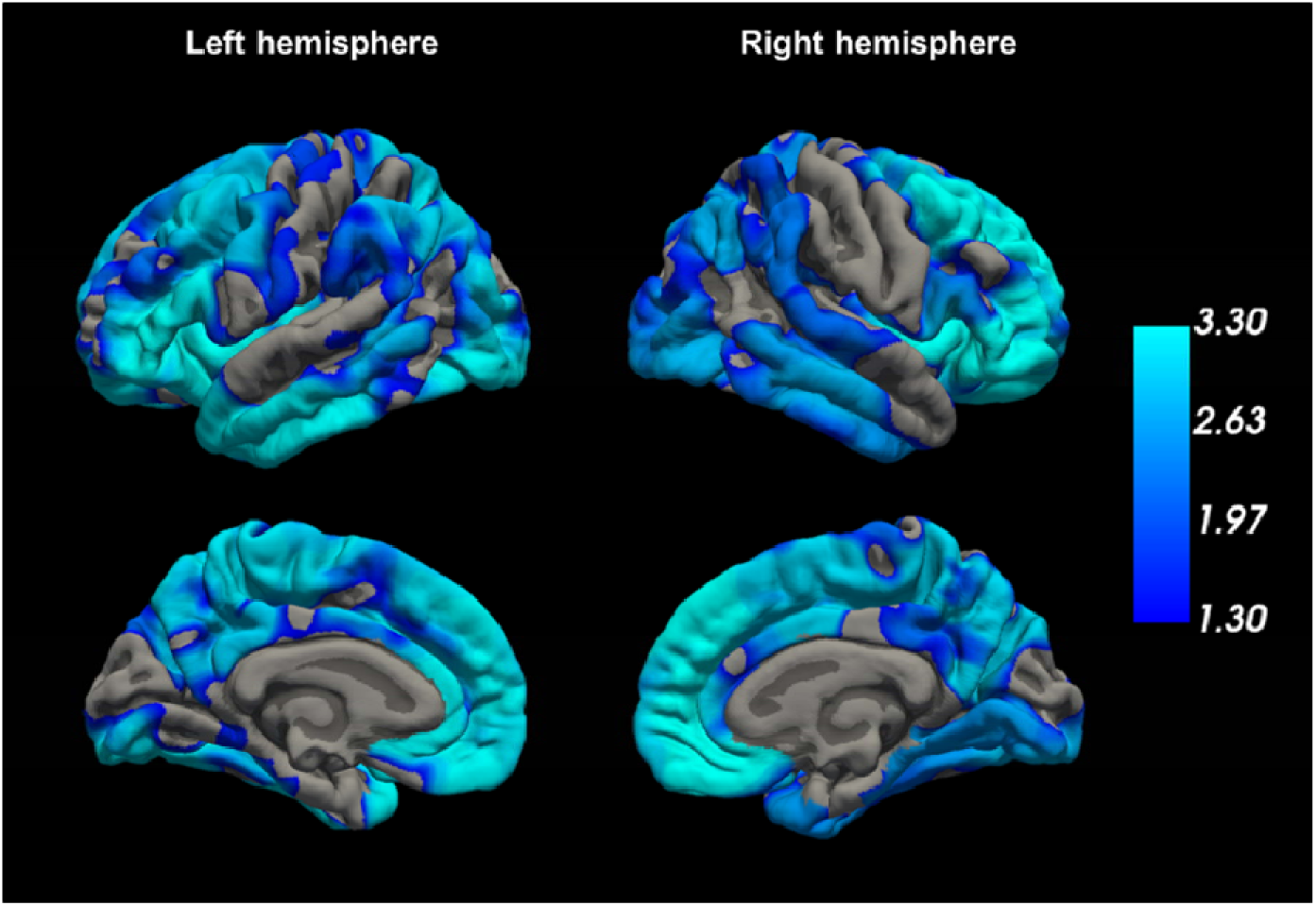
Cross-sectional associations between total ADHD scores and cortical surface area (Wave 1). Vertices with a negative association between total ADHD scores and surface area, projected onto the pial surface of the fsaverage brain template. There were no positive associations between ADHD scores and surface area. The color bar represents -log_10_(p) values. Displayed are all vertices with a TFCE-corrected p_FWE_<.05 (i.e. -log_10_(p)>1.3); grey – no significant association signal.

#### Secondary and sensitivity analyses

##### Global brain measures

Total ADHD scores were negatively related with total intracranial volume (vstat=-3.75, p=.001, df=10.78) and total surface area (vstat=-4.04, p=.0004, df=10.56), but not with mean cortical thickness (vstat=-0.32, p=.76, df=10.37). Following sensitivity analyses controlling for total surface area, the negative associations between total ADHD scores and the left and right hemisphere surface area clusters remained significant (left: vstat=-7.63, p=.0002, df=10.91; right: vstat=-6.31, p=.0002, df=10.91). When total intracranial volume was included as covariate, none of the subcortical volume associations remained significant.

##### Group comparisons

No significant diagnostic group differences were observed in the identified surface area clusters or in the volumes of the cerebellum, hippocampus, or amygdala.

##### Symptom domain-specific analyses

Models incorporating scores for inattention and hyperactivity-impulsivity separately yielded generally similar findings to those using total ADHD scores. Both inattention and hyperactivity-impulsivity scores were negatively related to surface area in one left-hemisphere and one right-hemisphere cluster, which were largely distributed over the cortical surface (Figure S1). No significant associations were found between either symptom score and cortical thickness. With regard to volumetric analyses, inattention was negatively associated with volumes of the right cerebellum (vstat=-3.02, p_FDR_=.02, df=10.62) and left amygdala (vstat=-3.25, p_FDR_=.02, df=10.72). Hyperactivity-impulsivity was negatively associated with volumes of left and right cerebellum (left: vstat=-2.84, p_FDR_=.02, df=9.96; right: vstat=-3.59, p_FDR_=.008, df=9.90), left and right caudate nucleus (left: vstat=-2.62, p_FDR_=.02, df=10.27; right: vstat=-2.73, p_FDR_=.008, df=10.44), right pallidum (vstat=-2.45, p_FDR_=.02, df=10.15), left and right hippocampus (left: vstat=-3.08, p_FDR_=.02, df=10.09; right: vstat=-2.74, p_FDR_=.008, df=10.03), and left and right amygdala (left: vstat=-3.81, p_FDR_=.02, df=10.08; right: vstat=-3.39, p_FDR_=.008, df=10.18).

### Longitudinal analyses

The distribution of total ADHD scores at each wave is shown in Figure 3. Additional plots showing the association between age and ADHD scores are provided Figure S2.

**Figure 3.**
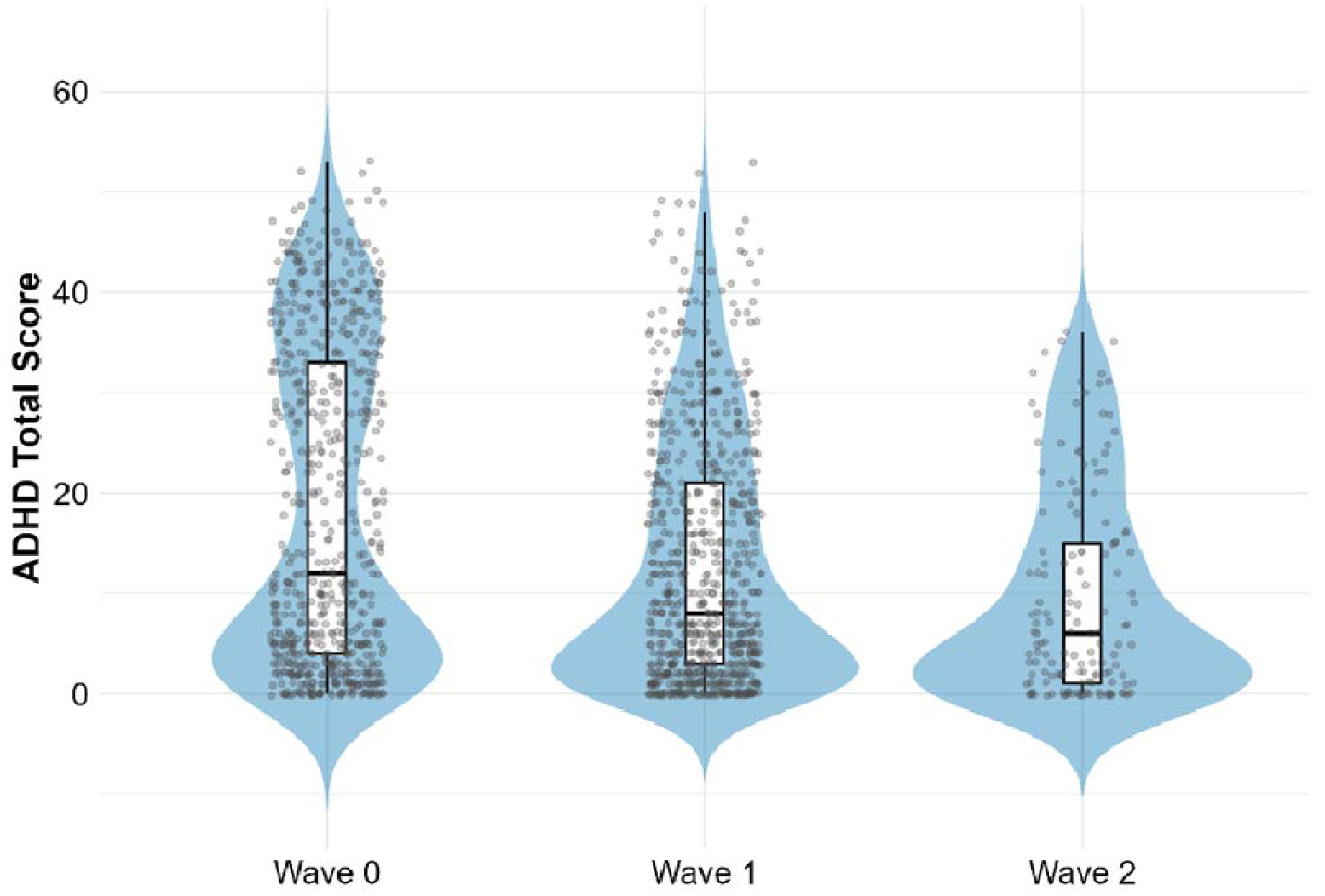
Distribution of ADHD total scores at Wave 0 (n=644), Wave 1 (n=765) and Wave 2 (n=149).

### Wave 0 to Wave 1

No significant associations were found between the change in ADHD scores (total or separate symptom domains) from Wave 0 to Wave 1 and any brain morphometric measures at Wave 1.

### Wave 1 to Wave 2

Between Wave 1 to Wave 2 a stronger decrease in total ADHD score was related to a stronger decrease or less increase in surface area in the left frontal pole/medial prefrontal cortex (size = 1455.46 mm^2^, p_FWE_<.05), and in the right occipital cortex (size=3935.80 mm^2^, p_FWE_<.05), right medial/orbitofrontal cortex (size=1319.72 mm^2^, p_FWE_ < .05), and right lateral orbitofrontal cortex (size=37.90 mm^2^, p_FWE_<.05; Figure 4a and Figure S3a). A stronger decrease in total ADHD score was related to a stronger decrease in cortical thickness in one large left-hemisphere and one large right-hemisphere cluster (left: size=37588.97 mm^2^, p_FWE_<.05; right: size=50468.70 mm^2^, p_FWE_<.05; Figure 4b and Figure S3b). No significant associations were found between change in total ADHD score and change in subcortical or cerebellar volumes. Furthermore, changes in total ADHD scores between Wave 1 and Wave 2 were not significantly associated with surface area, cortical thickness, or subcortical or cerebellar volumes measured at either Wave 1 or Wave 2. Surface area and mean cortical thickness values of the significant clusters in relation to age are displayed in Figure S4.

**Figure 4.**
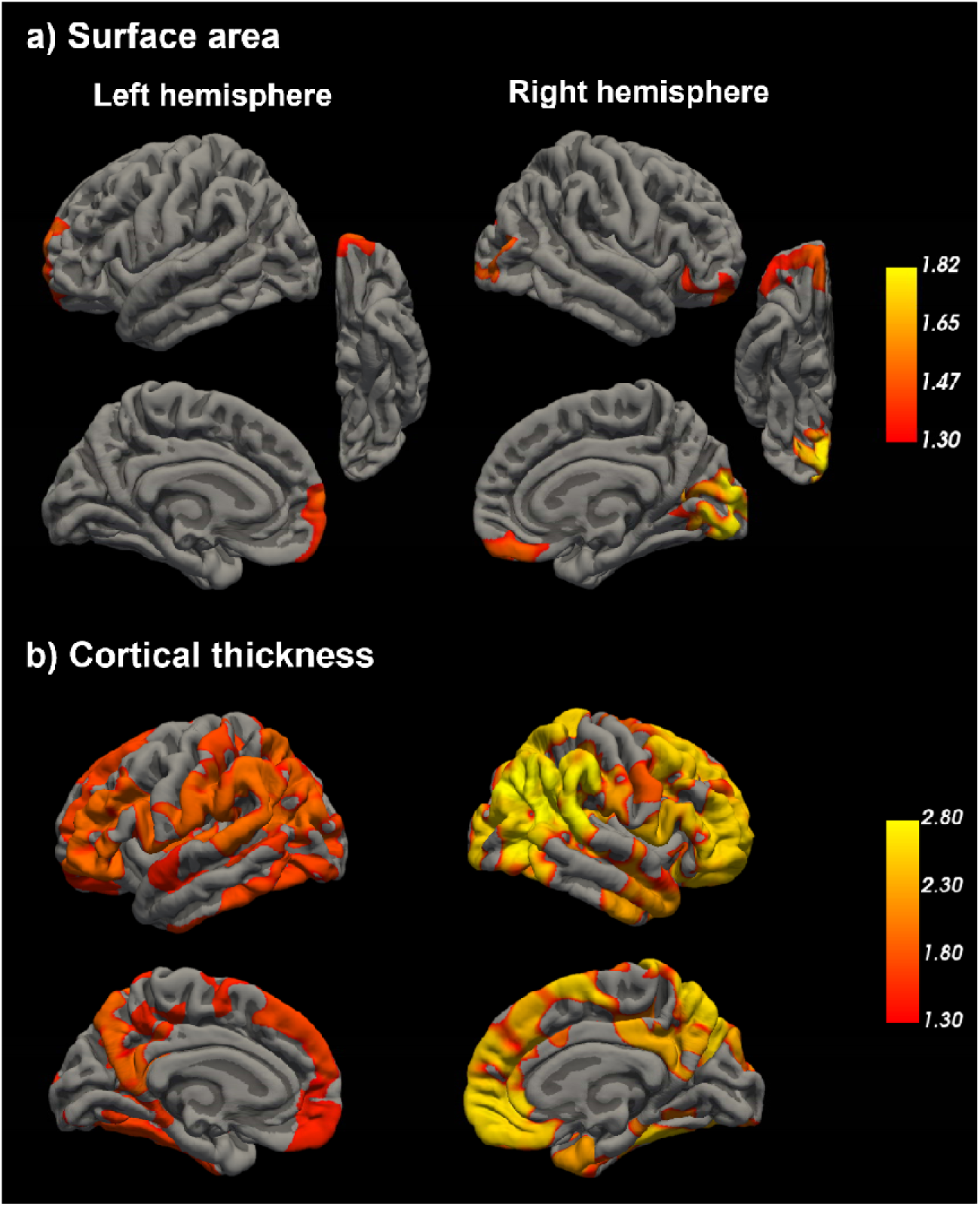
Longitudinal associations between change in total ADHD symptom scores and change in cortical morphometry (Wave 1 to Wave 2). Vertices with a positive association between change in total ADHD scores and change in a) surface area and b) cortical thickness between Wave 1 and 2, projected onto the pial surface of the fsaverage brain template. There were no negative associations between change in ADHD scores and change in surface area or cortical thickness. The color bar represents -log_10_(p) values. Displayed are all vertices with a TFCE-corrected p_FWE_<.05 (i.e. -log_10_(p)>1.3); grey – no significant association signal.

#### Secondary and sensitivity analyses

##### Global brain measures

Change in total ADHD score was positively related to both change in total surface area (vstat=2.67, p=.01, df=3.15) and change in mean cortical thickness (vstat=3.18, p=.003, df=3.06) between Wave 1 and Wave 2. Additionally, change in total ADHD score was positively associated with total intracranial volume at Wave 1 (vstat=2.32, p=.02, df=3.19) and Wave 2 (vstat=2.32, p=.02, df=3.25). Total surface area and mean cortical thickness values in relation to age are displayed in Figure S5. In sensitivity analyses including change in total surface area or mean cortical thickness as additional covariates, all original clusters showing positive associations with change in total ADHD scores remained significant.

##### Medication usage

When including medication usage as covariate, all clusters showing positive associations with change in total ADHD scores remained significant.

##### Symptom domain-specific analyses

Testing for inattention or hyperactivity-impulsivity symptoms separately as predictor variables showed that only change in inattention, but not change in hyperactivity-impulsivity, was positively associated with change in surface area in five clusters of the right hemisphere, which were located in prefrontal and occipital cortices (Figure S6). Changes in cortical thickness were related to changes in both symptom domains. Changes in inattention were positively associated with cortical thickness changes in one large left-hemisphere and one large right-hemisphere cluster (Figure S7a). Changes in hyperactivity-impulsivity were associated with cortical thickness changes in six left-hemisphere and two right-hemisphere clusters, widely distributed, including many prefrontal regions (Figure S7b). No significant associations were found between changes in inattention or hyperactivity-impulsivity scores and changes in subcortical or cerebellar volumes. Changes in hyperactivity-impulsivity were associated with surface area at Wave 1 in one cluster of the right hemisphere, peaking in the frontal pole. No other associations with brain morphometric measures at either Wave 1 or Wave 2 were observed.

##### Association with baseline ADHD scores

Higher Wave 1 symptom scores were associated with greater symptom reduction by Wave 2 (Figure S8). This prompted sensitivity analyses into potential impact of this collinearity between our predictors on the results. No significant associations between baseline ADHD scores and brain structural changes were observed (Figure S9) and the primary associations between changes in ADHD symptoms and brain structure remained significant when excluding baseline ADHD scores as covariate from the models.

## Discussion

This study investigated the developmental course of vertex-wise cortical thickness and surface area, and subcortical volumes in relation to ADHD symptom trajectories, including three waves of data collection, of which the last two included brain scans. On a cross-sectional level (Wave 1), higher ADHD symptoms were related to small reductions in surface area in many regions distributed over the cortical surface, including the frontal cortex, as well as to reductions in volumes of the cerebellum, amygdala, and hippocampus. In the longitudinal analysis from Wave 1 to Wave 2, a stronger decrease in ADHD symptoms was related to a stronger decrease in cortical thickness in many brain regions, and a stronger decrease or less increase in surface area in mainly frontal regions.

### Cross-sectional associations between ADHD symptom scores and brain morphometric measures

At Wave 1, higher ADHD symptom scores were associated with subtle but widespread reductions in cortical surface area, with strongest effects in the frontal cortex. Higher ADHD scores were also related to less total surface area. The identified brain regions remained significant after adjusting for total surface area, suggesting that these structural variations represent relatively larger effects in the identified cluster, relative to global measures or other brain regions. Our results align with previous studies showing reductions in surface area in relation to ADHD symptoms^5,29^, and highlight the importance of widespread cortical involvement beyond isolated prefrontal regions. In contrast to surface area, we found no significant cross-sectional associations of ADHD symptoms with cortical thickness at Wave 1. Those findings are in contrast to our hypotheses, but they are in line with previous literature showing more pronounced and stable associations between ADHD and surface area compared to cortical thickness^5,29^. In addition, more ADHD symptoms were associated with smaller volumes of the cerebellum, hippocampus, and amygdala, which is in line with previous cross-sectional studies^4^. However, these effects disappeared after controlling for total intracranial volume, which was also negatively related to ADHD scores, suggesting that subcortical reductions in this cohort may be part of more generalized, brain-wide variations in size rather than localized, region-specific effects.

### Longitudinal changes in brain morphometric measures related to changes in ADHD symptoms

Between Wave 1 and Wave 2, a stronger decrease in ADHD symptom scores was associated with a greater magnitude of cortical thinning across many brain regions. These effects appeared as large clusters in both hemispheres, encompassing frontal and parietal regions, as well as temporal and occipital areas. Symptom improvement was also associated with a greater reduction (or smaller increase) in surface area. In contrast to the widespread thinning, surface area changes were more localized to frontal areas and one occipital cluster. While change in ADHD symptoms was also associated with changes in mean cortical thickness and total surface area, the specific brain regions identified remained significant after controlling for these global measures, indicating a localized neurobiological process associated with symptom remission.

The direction of associations - symptom remission linked to brain "contraction" - may seem counter-intuitive given the cross-sectional finding of smaller surface area linked to more ADHD symptoms, and is also not in line with our initial hypothesis of longitudinal grey matter increases. However, it may be explained by the developmental window captured in our sample (Wave 1: mean age=16.45, SD=3.44 years; Wave 2: mean age=20.11, SD=3.47 years). Much of our current understanding of brain development in ADHD is derived from younger cohorts. The studies by Shaw et al.^11,12^ suggested a maturational delay in ADHD, characterized by reduced cortical thickness and surface area during childhood. While these differences converged with neurotypical peers during development, individuals with ADHD reached their maximal cortical expansion at a later age. Notably, the samples in the studies by Shaw were between eight and seventeen years old on average, with few participants in the older age range. In contrast, the period of adolescence into young adulthood captured in our study is characterized by different neurobiological processes.

During typical brain development, cortical thickness and surface area increase throughout childhood before peaking and subsequently declining throughout adolescence^12,30,31^. This reduction is driven by synaptic pruning and increased intracortical myelination, both of which refine neural circuits to enhance processing efficiency. Notably, cortical thickness and surface area follow distinct trajectories, with surface area peaking later^12,30,32^ and after that showing steady but more subtle decreases than cortical thickness^33^. Our findings align with these distinct patterns. The cross-sectional association between smaller surface area and higher ADHD symptoms at Wave 1 suggests that in adolescence those with higher symptoms may still be lagging in cortical expansion or just beginning their contraction phase. Meanwhile, the lack of cross-sectional cortical thickness findings suggests that the window for observing ADHD-related thickness reductions may have already passed in most individuals, as cortical thickness peaks earlier in development. Within this framework, our longitudinal results may reflect an adaptive maturational process or a maturational catch-up. If individuals with higher ADHD symptoms reach peak cortical expansion later, as the maturational delay theory suggests, then their subsequent phase of cortical thinning and surface area contraction also starts later. Our data suggest that in individuals experiencing the strongest symptom remission, this contraction phase may happen more rapidly or intensely. This supports the normalization theory of ADHD, where symptom reduction is biologically mirrored by a catch-up in cortical maturation. However, interpretations are limited by the sample’s wide age range, which spans multiple developmental stages within a single wave. Additionally, because imaging began at Wave 1 (average age 16), the lack of a "baseline" scan during early childhood (Wave 0) prevents us from fully mapping the earlier stages of the maturational delay hypothesis.

Concurrently, our findings may represent neural reorganization and compensation. The accelerated thinning and surface area contraction linked to symptom improvements may reflect a late-stage refinement of neural circuits that facilitates more efficient cognitive processing. Those with greater symptom improvement may have developed more effective coping or compensation mechanisms, which are reflected in more rapid or stronger cortical thinning and surface area contraction. This interpretation is supported by research into other domains of cognitive development. In neurotypical populations, higher intelligence has been linked more closely to the rate of structural change than to baseline morphometry^34,35^. Shaw et al.^34^ found that while children with a high IQ had a thinner cortex at age ten, they exhibited a thicker cortex in adolescence and a subsequent accelerated thinning in late adolescence, particularly in frontal regions. Similarly, Schnack et al.^35^ reported that once maximal values are reached, individuals with higher intelligence show faster thinning and surface area contraction. They suggested that this pattern might be explained by more efficient use of neural resources and increasing efficiency of structural networks over development. Our results tentatively suggest a similar mechanism in ADHD, where the trajectory of cortical change serves as a neurobiological marker of clinical improvement.

### Symptom domain-specific, global measures and subcortical patterns

We observed some symptom domain-specific trajectories. While changes in both inattention and hyperactivity-impulsivity were linked to widespread changes in cortical thickness, surface area changes in prefrontal and occipital clusters were primarily driven by improvements in inattention. Notably, while the rate of change was coupled between brain and behavior, baseline surface area and cortical thickness did not predict future symptom trajectories, nor did baseline symptoms predict future brain changes. In contrast, total intracranial volume stayed largely stable between Waves 1 and 2, but greater total intracranial volume both at baseline and follow-up was associated with a larger decrease in ADHD symptoms. Furthermore, despite finding cross-sectional subcortical associations, we found no evidence that subcortical or cerebellar volumes were associated with symptom changes over time. These results align with the neurodevelopmental model proposed by Halperin and Schulz^36^, which posits that while subcortical structures are involved in the early etiology of ADHD, cortical regions might drive symptom remission through compensatory maturation during the transition from adolescence to adulthood.

### Strengths and limitations

A major strength of this study is the use of one of the largest longitudinal datasets of children with ADHD to date, including a wide spectrum of ADHD symptom severities and trajectories. By utilizing a dimensional approach and permutation-based TFCE, we were able to capture the neurobiological spectrum of ADHD with enhanced sensitivity. The fact that significant associations emerged using dimensional scores, but not binary diagnostic categories, further supports the view that a dimensional approach may be more sensitive to capturing neurobiological correlates of ADHD^19,20^.

In addition to strengths, several limitations must be noted. First, the lack of a brain scan during early childhood and the large age variance within each wave should be considered when interpreting developmental trajectories. Additionally, while TFCE is highly sensitive to small but widespread effects and provides robust control for false positives, it offers less precision in localizing findings to very specific anatomical sub-regions. Nonetheless, given the distributed nature of ADHD-related brain changes, those widespread effects are likely more representative of the underlying neurobiology than highly localized effects.

## Conclusion

This study indicates that ADHD symptom trajectories are linked to patterns of cortical maturation during the transition from adolescence to adulthood. While lower surface area was a cross-sectional correlate of higher ADHD symptoms, longitudinal symptom remission was associated with accelerated cortical thinning and surface area contraction. These findings support a neural reorganization model of ADHD remission during development, where clinical improvement may be facilitated by a later-stage “catch-up” in cortical development.

## Data Availability

All data produced in the present study are available upon reasonable request to the authors

## Acknowledgements

The authors would like to thank all families who participated in this study and the researchers who collected the data. This study was funded by the European Union under the Marie Skłodowska-Curie grant agreement number 101065172. The data of this study was collected within the IMAGE and NeuroIMAGE projetcs. Those projects were supported by a Dutch Research Council (NWO) Large Investment Grant (no. 1750102007010) and NWO Brain & Cognition an Integrative Approach Grant (no. 433–09-242 to J.K.B.), and grants from Radboud University Medical Center, University Medical Center Groningen and Accare, and VU University Amsterdam. AW receives funding from the US National Institutes of Health (R01-MH139547, U54-HG013247, and R01-MH138425). BF acknowledges relevant funding by the Dutch Ministry of Education, Culture and Science of the government of The Netherlands for the NWO Gravitation programme GUTS (grant 024.005.011). Funding agencies had no role in study design, data collection, interpretation or influence on writing. Views and opinions expressed are those of the authors only and do not necessarily reflect those of the European Union or other funding agencies. Neither the European Union nor the granting authority can be held responsible for them.

## Author Contributions

Conceptualization and methodology: AM, ES. Software: AW. Resources: DvR, JO, CAH, PJH, ML, BF, JKB, ES. Data curation: AM, JK, AMS, DvR. Data analysis: AM. Supervision: ES. Writing draft: AM. Revision and editing of manuscript: All authors.

