## Supplement 1 for "ADHD symptom trajectories and brain morphometry: A longitudinal analysis"

**Structural MRI acquisition**

Structural MRI data in Wave 1 were collected on a Siemens MAGNETOM Avanto 1.5 Tesla system with 8-channel head coil in Nijmegen and on a Siemens MAGNETOM Sonata 1.5 Tesla system with the same 8-channel head coil in Amsterdam. In Wave 2, only participants from Nijmegen were invited and were scanned on the same scanner with the same head coil as in Wave 1. T1-weighted anatomical scans were acquired using a 3D magnetization prepared rapid acquisition with gradient echoes (MPRAGE) sequence (176 slices, flip angle=7°, TR=2730ms, TE=2.95ms, TI =1000ms, voxel size=1×1×1 mm, matrix size=256×256, parallel acquisition [GRAPPA] with an acceleration factor of 2). The same scanning protocol was used across scanners and waves.
