## Supplementary Figures for "ADHD symptom trajectories and brain morphometry: A longitudinal analysis"

a) **Inattention**

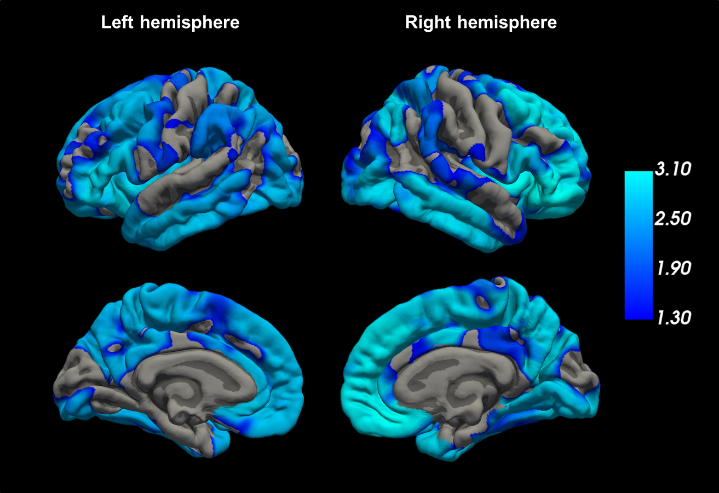

Left hemisphere cluster:

size = 54932.15 mm^2^

Right-hemisphere cluster:

size = 57229.90 mm^2^

b) **Hyperactivity-impulsivity**

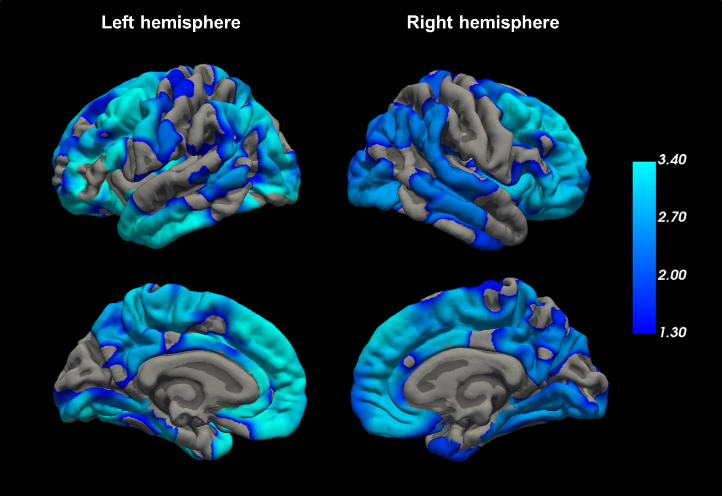

Left hemisphere cluster:

size = 53931.58 mm^2^

Right-hemisphere cluster:

size = 53118.44 mm^2^

**Figure S1.** **Cross-sectional associations between a) inattention and b) hyperactivity-impulsivity scores and cortical surface area (Wave 1).** Vertices with a negative association between a) inattention or b) hyperactivity-impulsivity scores and surface area, projected onto the pial surface of the fsaverage brain template. There were no positive associations between ADHD scores and surface area. The color bar represents -log_10_(p) values. Displayed are all vertices with a TFCE-corrected p_FWE_<.05 (i.e. -log_10_(p)>1.3); grey – no significant association signal.

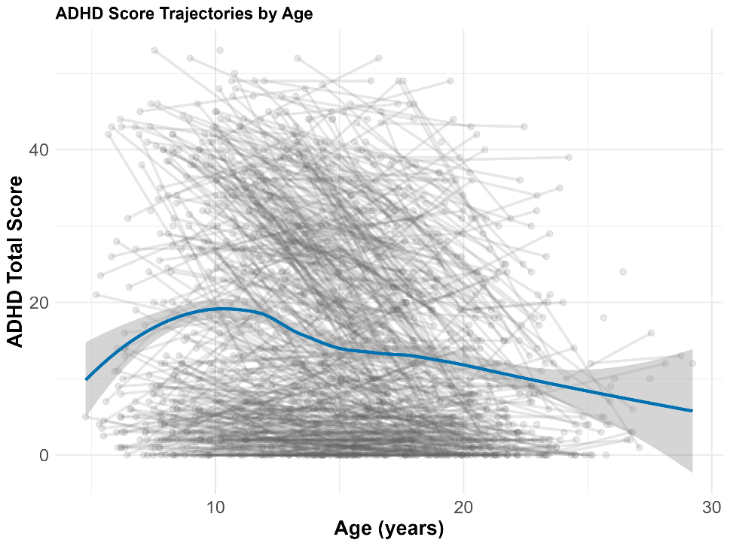

**Figure S2. Association between age and total ADHD scores including data from Wave 0 (n=644), Wave 1 (n=765) and Wave 2 (n=149).**

a) **Surface area**

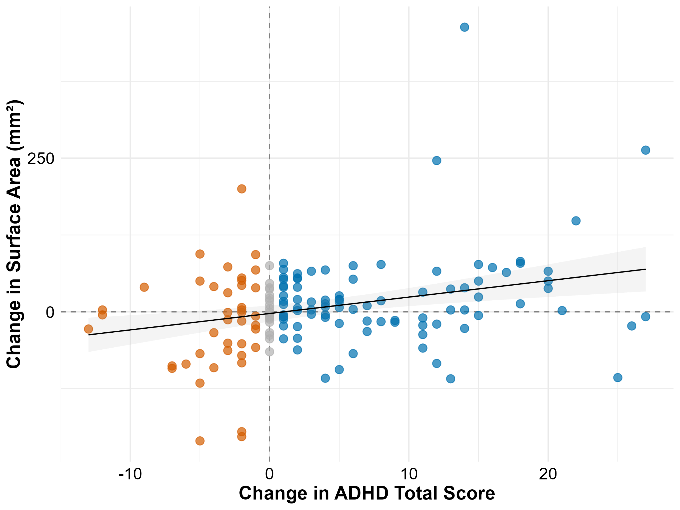

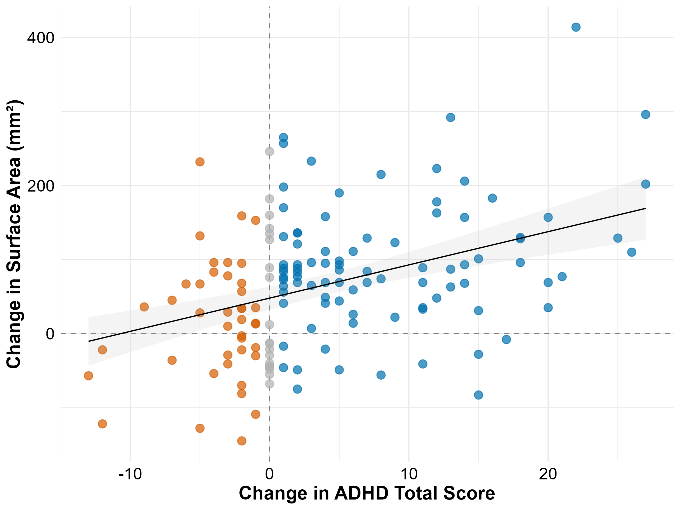

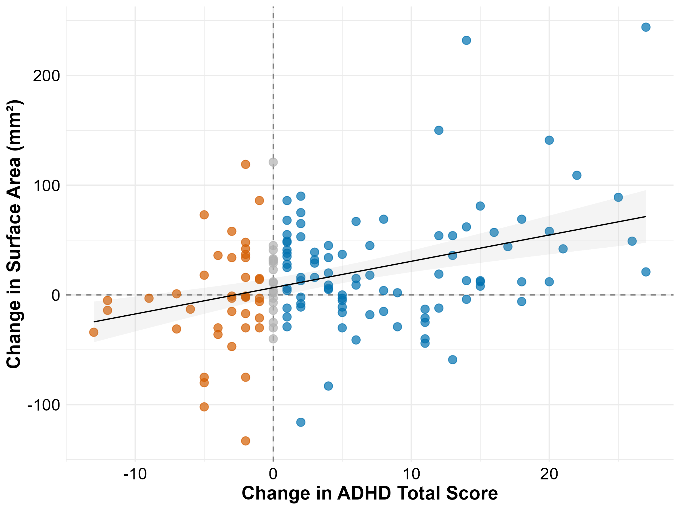
 **Left-hemisphere cluster	Right-hemisphere cluster 1**

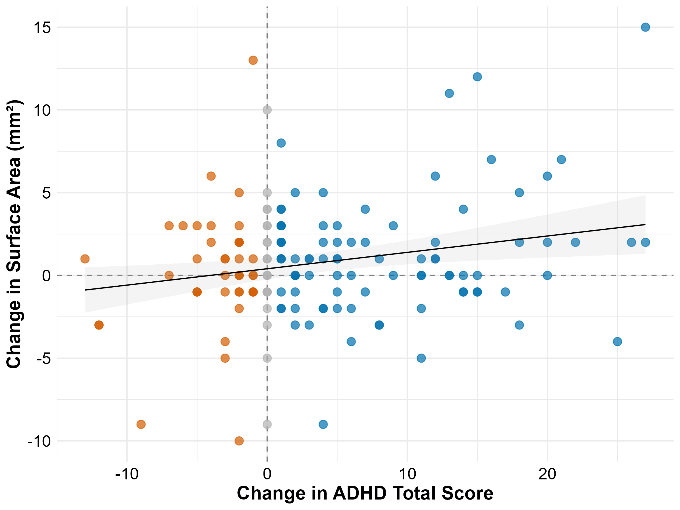
**Right-hemisphere cluster 2	Right-hemisphere cluster 2**

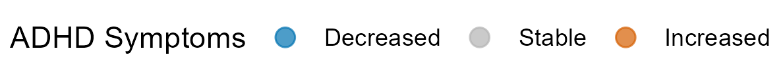

b) **Cortical thickness**

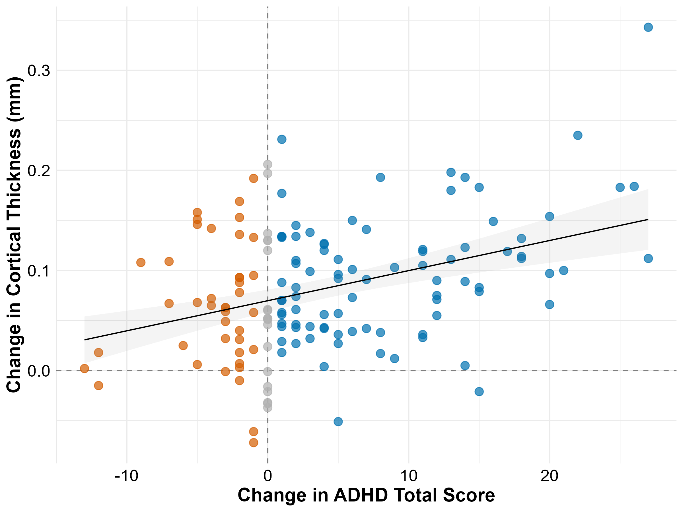

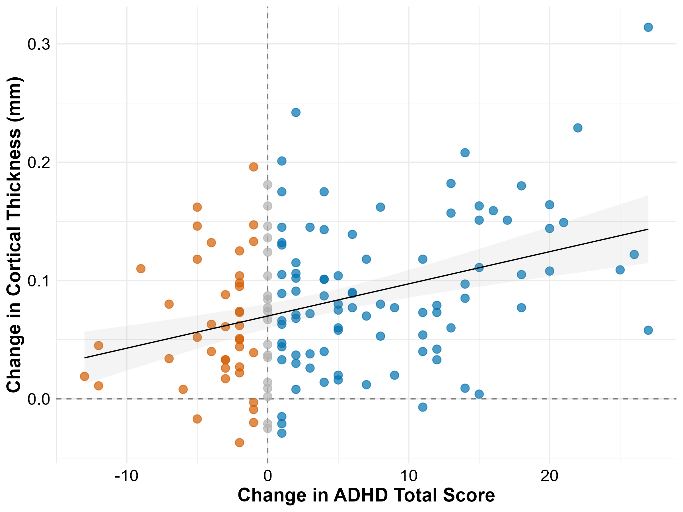
**Left-hemisphere cluster	Right-hemisphere cluster**

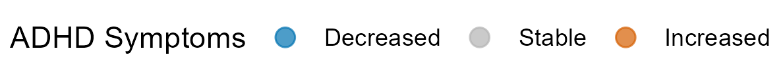

**Figure S3.** **Association between change in ADHD total score and change in a) surface area and b) cortical thickness in the significant clusters from Wave 1 to Wave 2.** Change was calculated as Wave 1 – Wave 2, so that more positive difference scores mean more decreases in ADHD score/brain morphometric measure and more negative values mean more increases in ADHD score/ brain morphometric measure.

a) **Surface area**

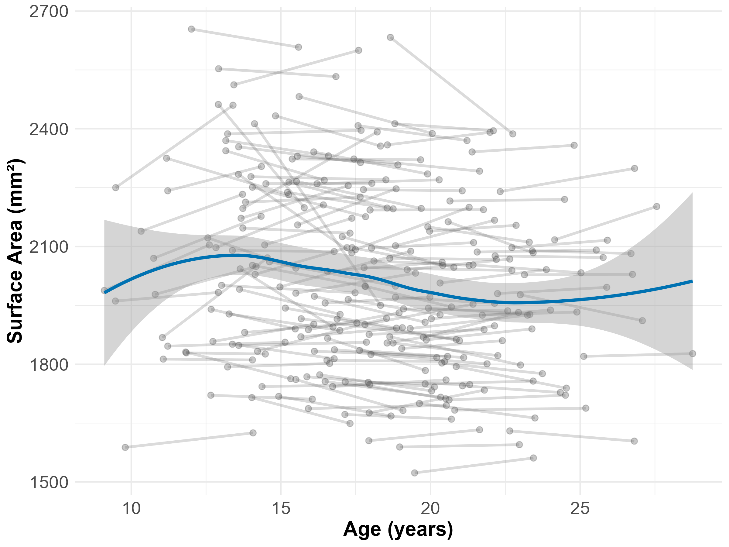
**Left-hemisphere cluster	Right-hemisphere cluster 1**

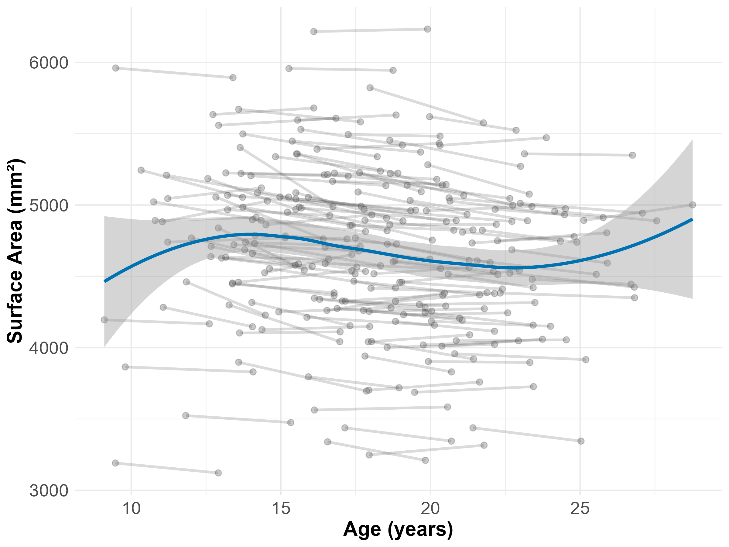

**Right-hemisphere cluster 2	Right-hemisphere cluster 3**

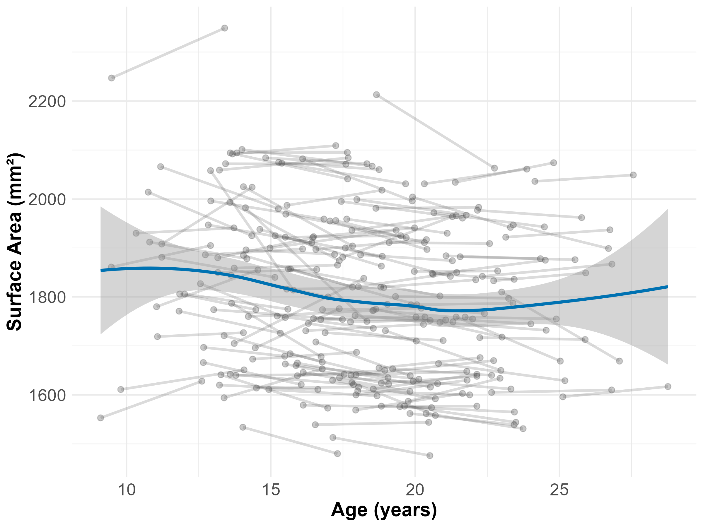

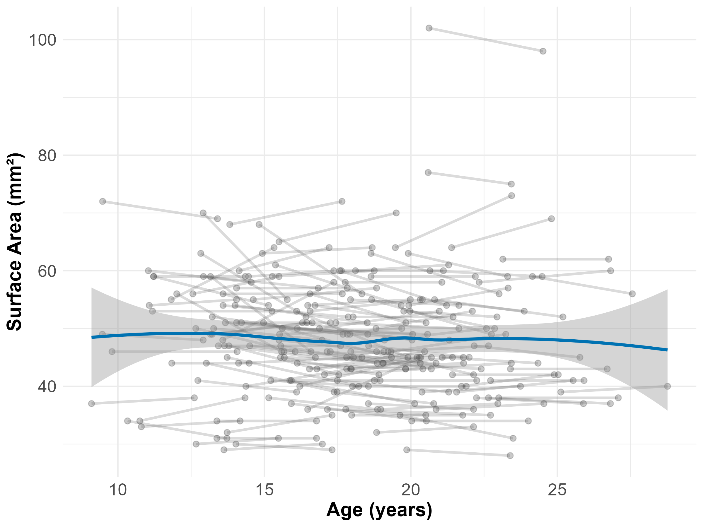

b) **Cortical thickness**

**Left-hemisphere cluster	Right-hemisphere cluster**

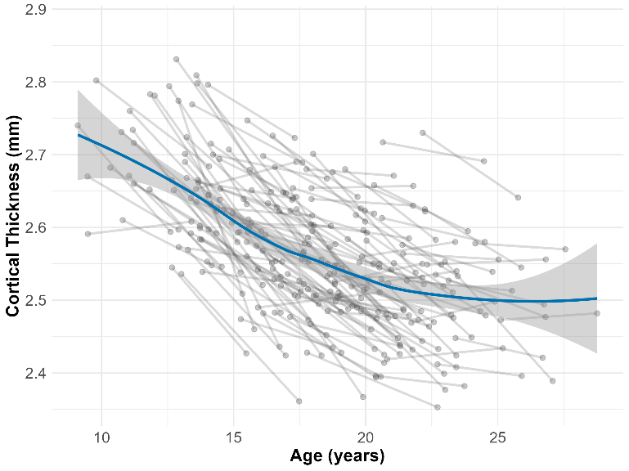

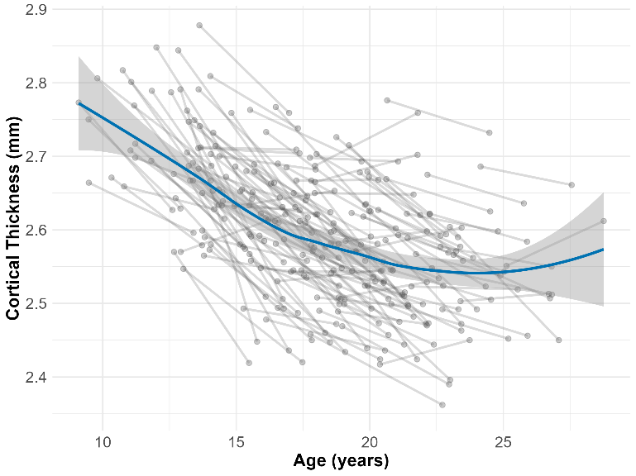

**Figure S4. Association between age and a) surface area and b) mean cortical thickness values of the significant clusters from the longitudinal analyses from Wave 1 to Wave 2.**

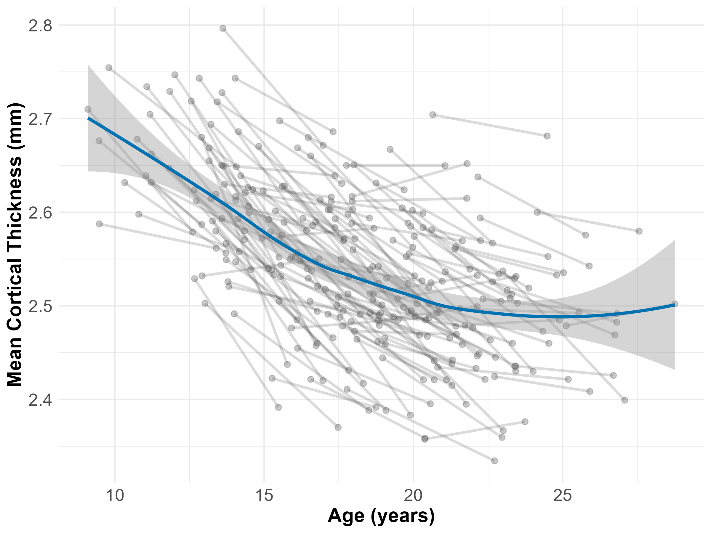

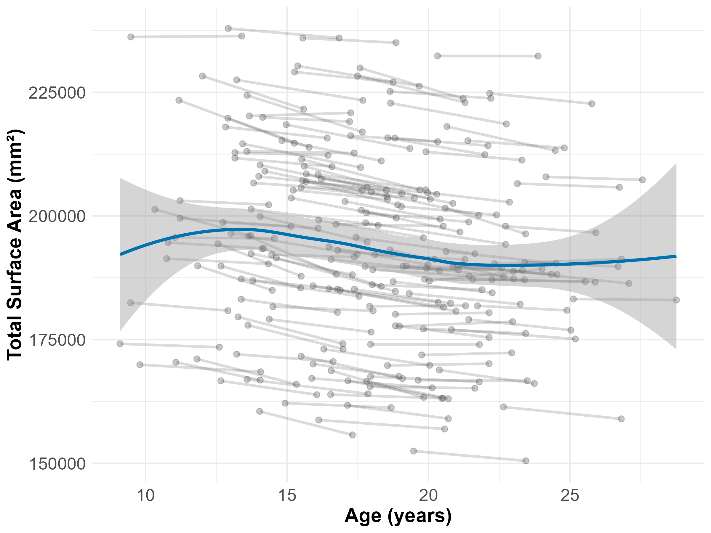

**Figure S5. Association between age and total surface area and mean cortical thickness including data from Wave 1 and Wave 2.**

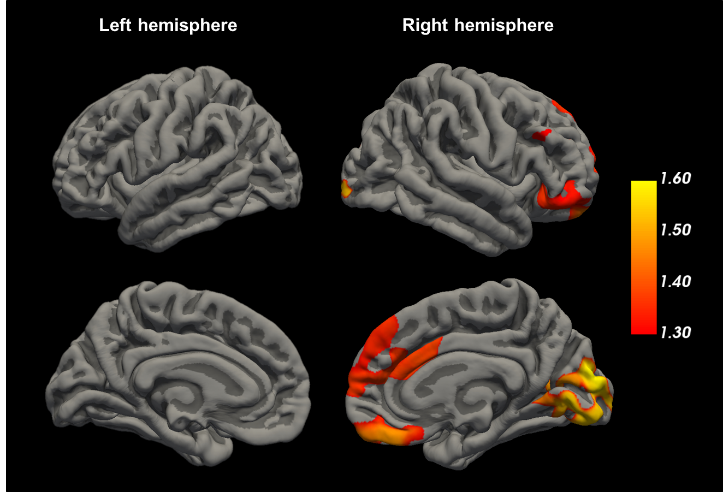

Left hemisphere: no significant cluster

Right-hemisphere clusters:

cluster 1: size = 3174.16 mm^2^

cluster 2: size = 1758.04 mm^2^

cluster 3: size = 1644.55mm^2^

cluster 4: size = 217.78 mm^2^

cluster 5: size = 79.94 mm^2^

**Figure S6. Longitudinal associations between change in inattention scores and change in cortical surface area (Wave 1 to Wave 2).** Vertices with a positive association between change in inattention scores and change in surface area between Wave 1 and 2, projected onto the pial surface of the fsaverage brain template. There were no negative associations between change in inattention scores and change in surface area. The color bar represents -log_10_(p) values. Displayed are all vertices with a TFCE-corrected p_FWE_<.05 (i.e. -log_10_(p)>1.3); grey – no significant association signal.

a) **Inattention**

Left hemisphere cluster:

size = 15992.24 mm^2^

Right-hemisphere cluster:

size = 48437.63 mm^2^

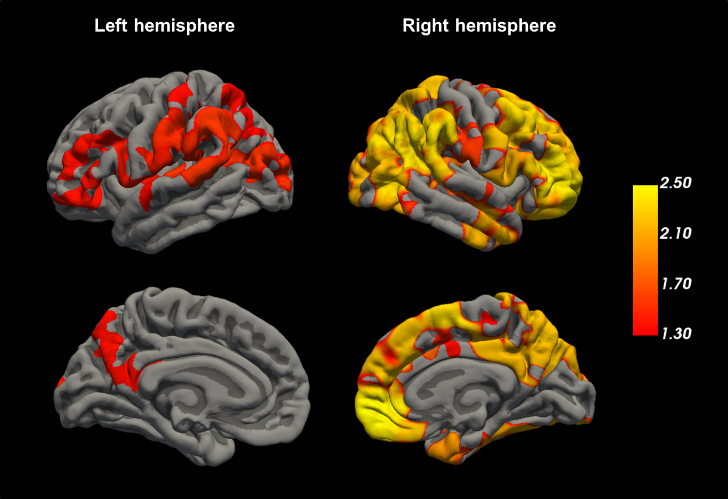

b) **Hyperactivity-impulsivity**

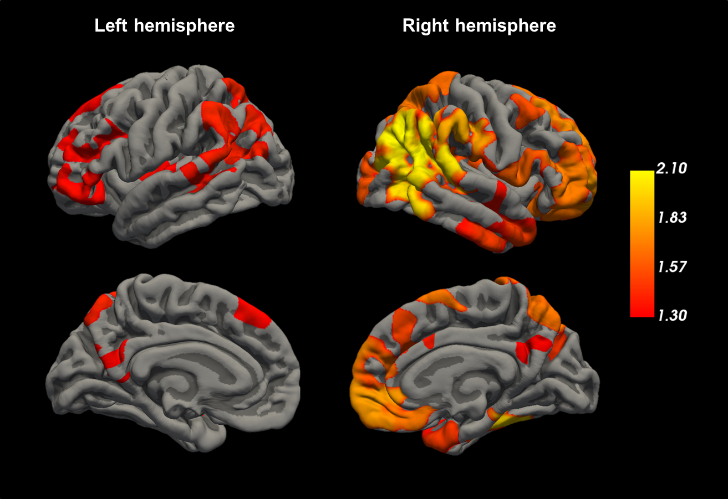

Left hemisphere clusters:

cluster 1: size = 6155.86 mm^2^

cluster 2: size = 1527.65 mm^2^

cluster 3: size = 1513.63 mm^2^

cluster 4: size = 1389.60 mm^2^

cluster 5: size = 878.81 mm^2^

cluster 6: size = 820.22 mm^2^

Right-hemisphere clusters:

cluster 1: size = 33239.52 mm^2^

cluster 2: size = 375.17 mm^2^

**Figure S7. Longitudinal associations between change in a) inattention scores and b) change in hyperactivity-impulsivity scores and change in cortical thickness (Wave 1 to Wave 2).** Vertices with a positive association between change in a) inattention or b) hyperactivity-impulsivity scores and change in cortical thickness between Wave 1 and 2, projected onto the pial surface of the fsaverage brain template. There were no negative associations between change in inattention or hyperactivity-impulsivity scores and change in cortical thickness. The color bar represents -log(p) values. Displayed are all vertices with a TFCE-corrected p_FWE_<.05 (i.e. -log_10_(p)>1.3); grey – no significant association signal.

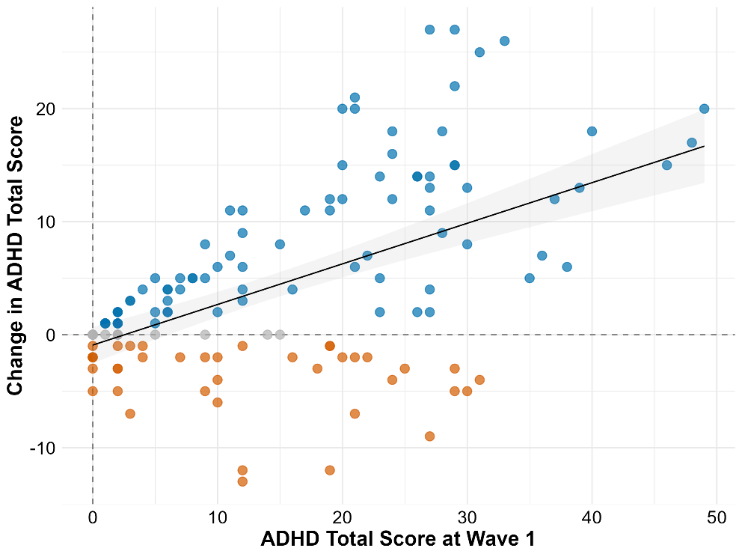

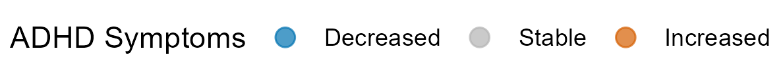

**Figure S8.** **Association between ADHD total score at Wave 1 and changes in ADHD total score from Wave 1 to Wave 2.** Change was calculated as Wave 1 – Wave 2, so that more positive difference scores mean more decreases in ADHD score and more negative values mean more increases in ADHD score.

a) **Surface area**

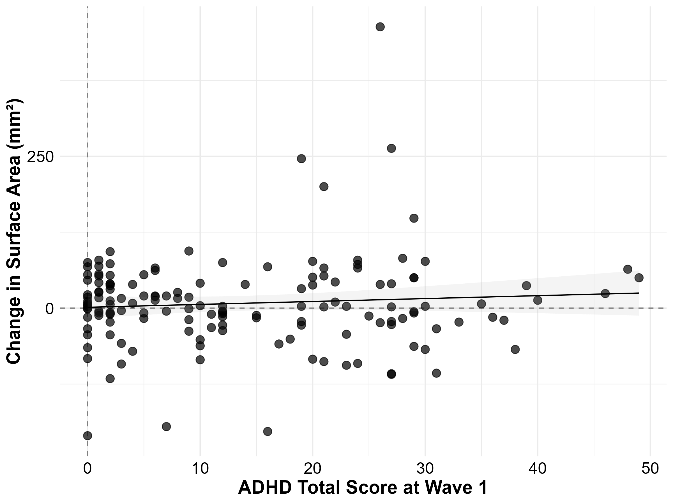

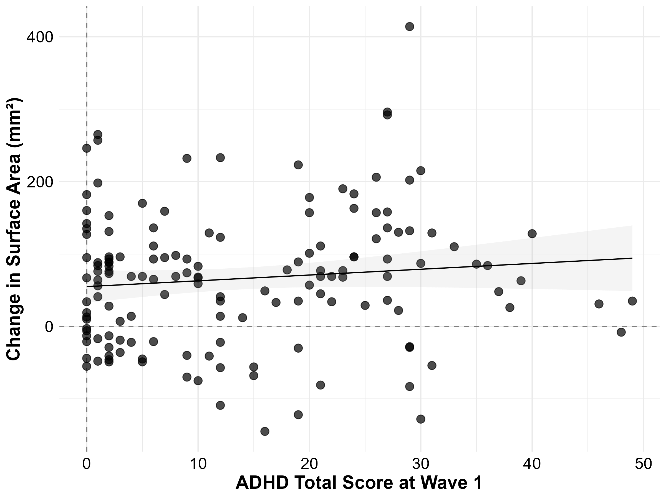
**Left-hemisphere cluster	Right-hemisphere cluster 1**

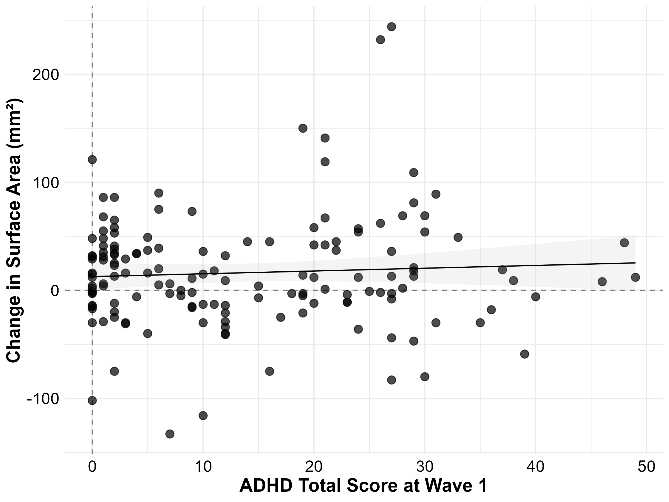

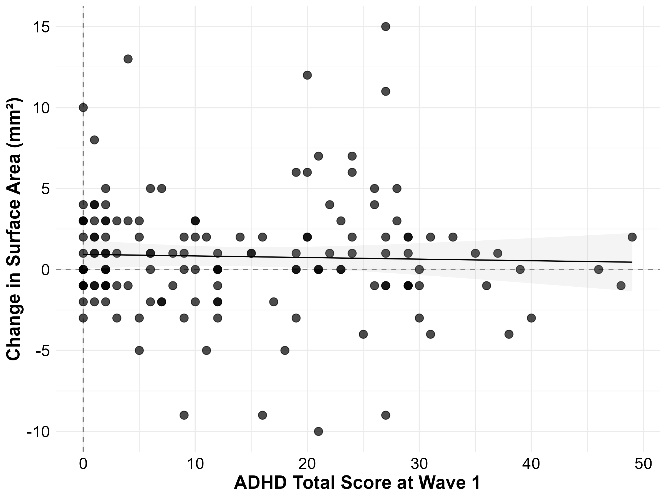
**Right-hemisphere cluster 2	Right-hemisphere cluster 3**

b) **Cortical thickness**

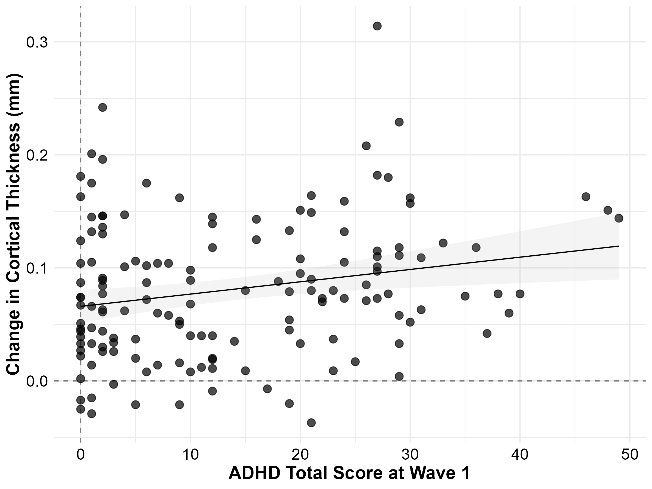

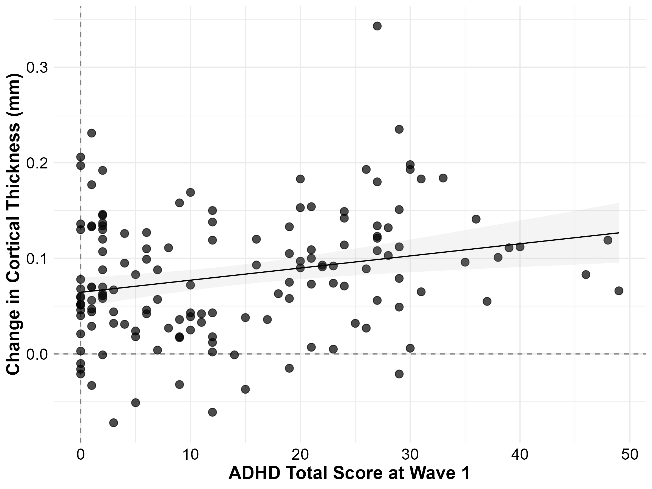
**Left-hemisphere cluster	Right-hemisphere cluster**

**Figure S9. Association between ADHD total score at Wave 1 and changes in a) surface area and b) cortical thickness in the significant clusters from Wave 1 to Wave 2.** Change was calculated as Wave 1 – Wave 2, so that more positive difference scores mean more decreases in surface area/cortical thickness and more negative values mean more increases in surface area/cortical thickness.
